# Discovery of Urinary Metabolite Biomarkers of Psychiatric Disorders Using Two-Sample Mendelian Randomization

**DOI:** 10.1101/2023.09.26.23296078

**Authors:** Jihan K. Zaki, Jakub Tomasik, Jade McCune, Oren A. Scherman, Sabine Bahn

## Abstract

**Background:** Psychiatric disorders cause substantial patient suffering world-wide, which could be alleviated through the discovery of early diagnostic biomarkers. Urinary markers have promising practical applications; however, no robust urine biomarkers exist currently for psychiatric disorders. While the traditional biomarker discovery process is costly and time-consuming, genetic methods utilizing existing data from large-scale studies, such as Mendelian randomization, may provide an alternative, cost-effective approach to identifying such biomarkers.

**Methods:** A two-sample Mendelian randomization analysis was conducted in R using GWAS data for seven psychiatric disorders from the Psychiatrics Genomics Consortium, as well as a meta-analysis of urinary metabolite GWAS studies and the GWAS Catalog. Mendelian randomization assumptions were assessed using the MR-Egger intercept, P-values, and genetic associations from the PhenoScanner database.

**Outcomes:** The Mendelian randomization analysis revealed 67 analyte-disorder associations, of which 21 were exclusive to a single disorder. Most notable associations were observed between tyrosine and schizophrenia (*β*=−0.041, SE=0.013, Q=0.027), and creatine and bipolar disorder (*β*=−0.077, SE=0.019, Q=0.002), which validated across multiple studies, as well as that of pyridoxal (*β*=0.10, SE=0.03, Q=0.042) and ferulic acid 4-sulfate (*β*=0.077, SE=0.025, Q=0.037) to anorexia nervosa, and N,N-dimethylglycine to attention deficit hyperactivity disorder (*β*=−0.39, SE=0.11, Q=0.008).

**Interpretation:** These results indicate an association between bipolar disorder, schizophrenia, anorexia nervosa, and attention deficit hyperactivity disorder with urinary metabolite marker alterations. Most of the findings were supported by previous literature. The results provide a roadmap for future experimental and clinical validation of the identified biomarker candidates and demonstrate the utility of using genetic instruments for urinary biomarker discovery.

## 1. Introduction

Psychiatric disorders pose significant diagnostic challenges, leading to substantial personal and economic burden worldwide. The 12 most debilitating psychiatric disorders are estimated to annually cause 125 million disability adjusted life years globally^1^, and economically the direct and indirect costs of mental health disorders in the UK alone are estimated to account for 4.5% of the gross domestic product, or £70B-£100B every year^2^. Only the three most debilitating psychiatric conditions, i.e., major depressive disorder, schizophrenia, and bipolar disorder, have each been estimated to incur annual societal costs of £5B-£8B in England alone^3–5^.

Although early intervention and treatment have been associated with improved long-term outcomes^6–8^, delays in the diagnosis of psychiatric disorders are common. The difficulty in diagnosing psychiatric diseases lies within the current diagnostic frameworks, which are primarily interview-based methods^9,10^, as well as the lack of biological consistency within the disorders^11^. As a result, 39% of patients with a severe psychiatric disorder are initially misdiagnosed^12^. It has been reported that the average diagnostic delay for bipolar disorder is 6.5 years^13^, and the mean duration of untreated psychosis has been estimated to be up to 2.9 years^14^. Early and accurate diagnosis and intervention could significantly mitigate the severe burden and costs associated with mental health disorders. This could be facilitated by the discovery of consistent and actionable biomarkers in easily accessible patient samples, which could additionally provide a valuable means to improve biological understanding of psychiatric diseases.

Among other biological patient samples, urine holds great promise as a potential source of biomarkers. However, it is commonly overlooked by the scientific community for nearly all disorders. Publications listed in PubMed for blood-based biomarker research outnumber urinary biomarker publications ten-fold. Despite being often overlooked, urine mostly retains the biomarker signal observed in plasma, as a substantial proportion of urinary metabolite concentrations are filtered directly from plasma^15^. Furthermore, urine has significant advantages as a biomarker source compared to other body fluids due to the ease and non-invasive nature of sample collection, and the abundance of sample volume. Discovery of robust urinary markers could introduce an opportunity for a simple, non-invasive, and longitudinal biomarker assessment as well as point-of-care testing for high risk of disease or relapse.

The high clinical potential of urinary biomarkers can be fully utilised with careful study design and appropriate methodology. Computational methods could overcome the challenges of urinary biomarker discovery in psychiatric diseases, including the high variation in analyte concentrations^16,17^ as well as the significant impact of potential confounding effects. One such method is a genetic analysis called Mendelian randomization (MR)^18^, which utilizes the principles of instrumental variable analysis and applies them to genetic data to evaluate the effects of a given exposure (i.e., an analyte of interest) on a selected outcome (i.e., risk of psychiatric disease) when the effect of the genetic variability on both are known.

In the present study, we aimed to discover genetically linked urinary markers for seven psychiatric disorders potentially capable of differential diagnosis. A two-sample MR study was conducted using exposure data obtained from a meta-analysis of urinary metabolite GWAS studies and urinary metabolite associations from the GWAS catalog, as well as outcome data obtained from the most up-to-date psychiatric disorder GWAS data. Additionally, sensitivity analyses were conducted, including tests for horizontal pleiotropy, heterogeneity, confounder associations, as well as the determination of potential mechanistic reasons for altered expressions, to validate MR assumptions and ensure the robustness of the identified biomarker candidates.

## 2. Methods

### 2.1. Compilation of the exposure data

The exposure association data were obtained from a meta-analysis of urinary metabolite GWAS described in detail by Zaki et al. (manuscript in preparation), as well as results from the GWAS catalog under the trait urinary metabolite measurement (EFO 0005116). In brief, the meta-analysis^19^ combined results from five urinary metabolite GWAS^20–24^ using a sample size-based meta-analysis method^25^, identifying 2248 significant associations for 14 analytes (P*<*7.1×10^−9^) prior to clumping. The identified SNPs from the meta-analysis were combined with 195 associations for 152 analytes obtained from the GWAS catalog^20,21,23,24,26–30^, excluding SNPs duplicated in the catalog as well as mislabelled studies conducted in other sample types. All studies contained European ancestry populations; however, some studies differed in confounder profiles such as obesity and age. Exclusively genome-wide significant associations (P*<*5×10^−8^) were selected for the analysis from the GWAS catalog. An F-statistic was calculated for all exposure associations to evaluate weak instrument bias^18^.

### 2.2. Psychiatric disorder GWAS data collection

The most recent summary statistics GWAS data for each assessed psychiatric disorder were selected from meta-analyses conducted by the Psychiatrics Genomics Consortium, which were primarily based on European ancestry populations and did not have patient overlap with the exposure datasets. The seven psychiatric disorders selected for this study included attention deficit hyperactivity disorder (n_ADHD_=20183, n_CTRL_=35191)^31^, anorexia nervosa (n_ANO_=16996, n_CTRL_=55525)^32^, autism spectrum condition (n_ASC_=18381, n_CTRL_=27969)^33^, bipolar disorder (n_BD_=41917, n_CTRL_=371549)^34^, major depressive disorder (n_MDD_=15771, n_CTRL_=178777)^3^5, schizophrenia (n_SCZ_=76755, n_CTRL_=243649)^36^, and Tourette’s syndrome (n_TS_=4819, n_CTRL_=9488)^37^.

### 2.3. Mendelian randomization

Two-sample MR was conducted to genetically assess causal associations of analytes selected in the meta-analysis and the GWAS catalog to the seven psychiatric disorders. MR analysis was carried out using the TwoSampleMR^38^ package in R. For the significant analyte-SNP associations from the meta analysis, MR was performed separately between each of the meta-analysed studies and the outcome datasets, with MR estimates subsequently combined using the inverse variance weighted (IVW) meta-analysis method^25^. This two-step process was necessary due to inconsistent measurement methods, scales, and log transformations used for individual analytes included in the meta-analysis, which caused their beta coefficients and standard errors to be incomparable and prevented them from being combined into a single coefficient. For the exposure data from the GWAS catalog, a standard single-step MR was performed.

For each of the analyses, closely located (1000 kb) and correlated (r^2^*<*0.001) SNPs in the exposure dataset were combined by clumping using the European population-based reference panel from the 1000 Genomes Project^39^. For exposure SNPs not found in the outcome dataset, highly correlated (r^2^*<*0.8) linkage disequilibrium (LD) proxy replacements in the outcome dataset were identified using the European population from the 1000 Genomes Project as a reference, and the LDlinkR^40^ package in R. The IVW ratio MR method was used for metabolites with at least two SNP associations, and the Wald ratio (WR) MR method was used for metabolites with only a single SNP association^41^. For metabolites with three or more SNP associations, sensitivity analyses were conducted using the weighted median, weighted mode, and MR-Egger methods to assess consistent directionality and effect sizes^42^. Cochrane’s Q test was applied for metabolites assessed using the inverse variance weighted method to assess heterogeneity between effect size estimates^42^. The Benjamini-Hochberg method was used to correct for multiple comparisons, with the significance threshold set to Q*<*0.05.

### 2.4. Assumption validation

The three assumptions of MR analysis state that: 1) the genetic instrument must be associated with the exposure, 2) the genetic instrument must not directly influence the outcome (i.e., horizontal pleiotropy must not occur), and 3) the genetic instrument must not be associated with potential confounders of the exposure and the outcome^18^. These assumptions were assessed for all results which passed the significance threshold following multiple comparison correction. The first assumption of genetic instrument association to exposure levels was assessed through the significance level of the exposure associations (P*<*5×10^−8^) in either the GWAS catalog or the meta-analysis. To assess the second and third assumptions, SNP-trait associations were examined using the PhenoScanner database^43,44^, with genome-wide significant associations to either the disorder or known demographical confounders (such as smoking for schizophrenia) invalidating the assumptions. Additionally, MR-Egger intercept was used to assess metabolites with more than three SNP associations, with significant associations indicating horizontal pleiotropy^42^.

## 3. Results

### 3.1. Mendelian randomization

In the present work, the causal effect of 163 urinary analytes on seven selected psychiatric disorders was assessed using two-sample MR, with a total of 4852 analyte-SNP associations combined from the meta-analysed studies (4657) and the GWAS catalog (195) in the exposure dataset, and 56,698,746 unique SNP measurements in the outcome dataset. Prior to assumption validation, 68 statistically significant metabolite-disorder associations were discovered between 45 unique analytes and the mental health conditions ADHD, anorexia, BD, and schizophrenia following multiple comparison adjustment.

### 3.2. Assumption validation and sensitivity analyses

To ensure that the first assumption of MR is met, and the instrumental variables are robustly associated with exposure, the analysis included only those SNP-analyte associations that were genome-wide significant (P*<*5×10^−8^) in either the GWAS catalog or the meta-analysis. The second and third assumption of MR was assessed using SNP-trait associations from the PhenoScanner database. The analysis revealed a genome-wide significant association of alcohol intake frequency with rs2287921 and rs281408, which were both significantly associated with fucose. Therefore, the association of fucose to schizophrenia (*β*=0.09, SE=0.017, Q=3.56×10^−5^) was excluded from the list of results, as potentially representing an indirect effect of alcohol consumption. No other analysed SNPs were found to violate the assumptions of MR. Furthermore, horizontal pleiotropy was not found to be present in any of the SNP-trait associations where an MR-Egger intercept was able to be calculated (≤3 SNPs per analyte). Further sensitivity analyses showed that for associations whose exposure data were obtained from multiple studies, the MR estimates were consistent between individual source studies, as shown in Supplementary Figures 1 and 2. Additionally, no heterogenic effects were observed for SNPs involved in any of the significant associations (Supplementary Table 2). Finally, consistent effect sizes were observed for all applicable SNP-trait associations using non-primary MR modes (weighted mode, weighted median, and MR-Egger) which have been presented in Supplementary Figure 3.

### 3.3. Common markers of psychiatric disorders

Following the assumption validation, 67 metabolite-disorder associations remained significant between 44 unique analytes and the respective disorders, with no significant associations found for ASC, MDD, or Tourette’s syndrome. All significant metabolite-disorder associations following meta-analysis and assumption validation are presented in Supplementary Table 1. The largest overlap was observed between biomarker associations to BD and schizophrenia, which overlapped in 22 unique analytes with similar effect sizes and directionality. The extent of overlap between BD and schizophrenia is shown in Figure 1. The most significant associations overlapping between BD and schizophrenia involved most notably numerous N-acetylated compounds. In addition, anorexia was associated with upregulated pyridoxal (*β*=0.10, SE=0.03, Q=0.042) and ferulic acid 4-sulfate (*β*=0.077, SE=0.025, Q=0.037), which also overlapped with BD and schizophrenia, however with opposite directionality.

**Figure 1:**
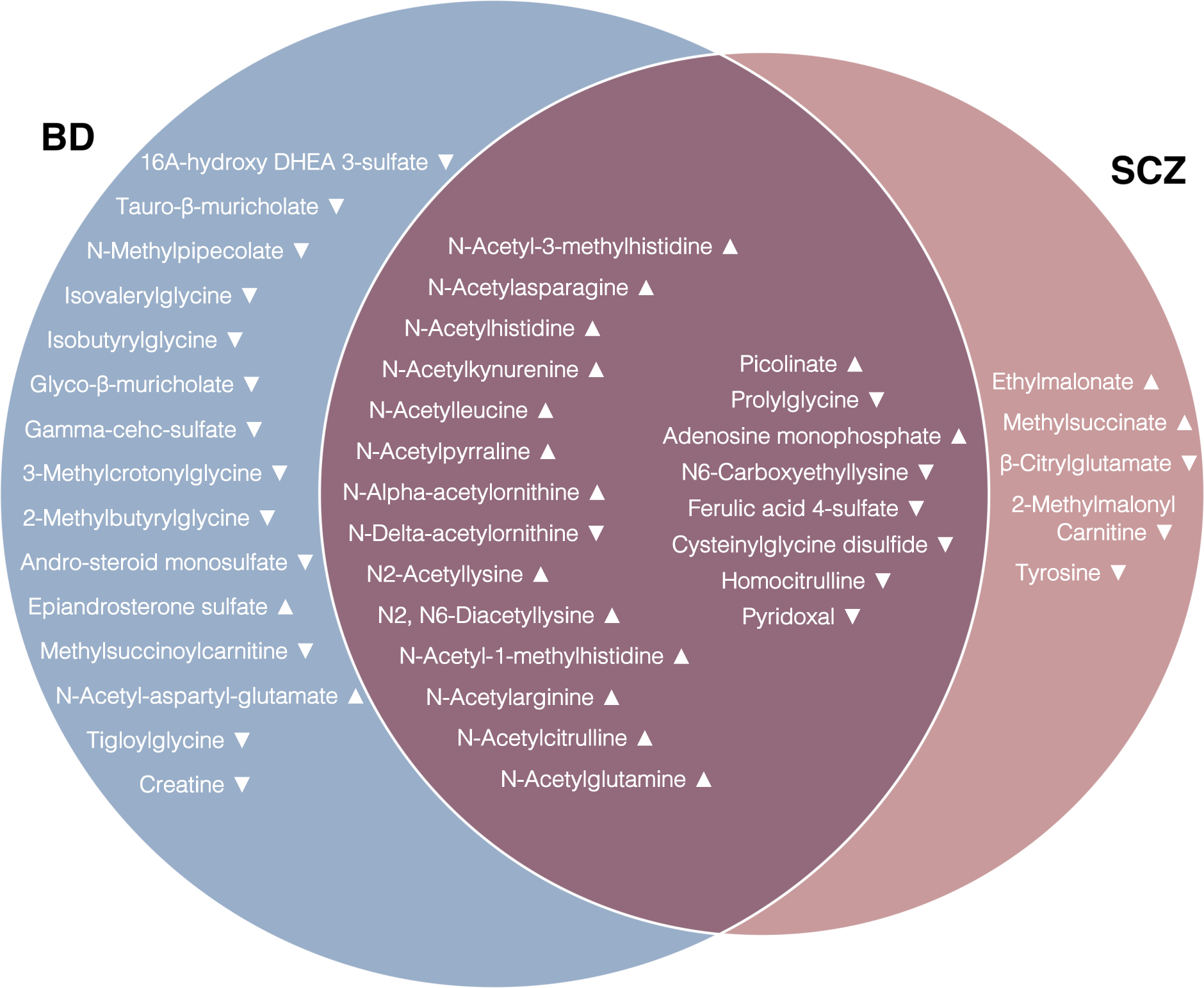
Venn diagram of significant analyte association for schizophrenia (SCZ), and bipolar disorder (BD). Effect size estimate directionality has been indicated with triangles.

### 3.4. Differential markers of psychiatric disorders

Of the 67 significant metabolite-disorder associations, 21 were exclusive to a single disorder. Of these, five were associated with schizophrenia, 15 with BD, and one with ADHD. The most robust associations were observed between tyrosine and schizophrenia (*β*=−0.041, SE=0.013, Q=0.027), and creatine and BD (*β*=−0.077, SE=0.019, Q=0.002), due to their associations being significant after assessment in multiple studies. The single analyte association to ADHD was N,N-dimethylglycine (*β*=−0.39, SE=0.11, Q=0.008). All non-overlapping analyte associations specific to schizophrenia or BD are shown in Figures 2.

**Figure 2:**
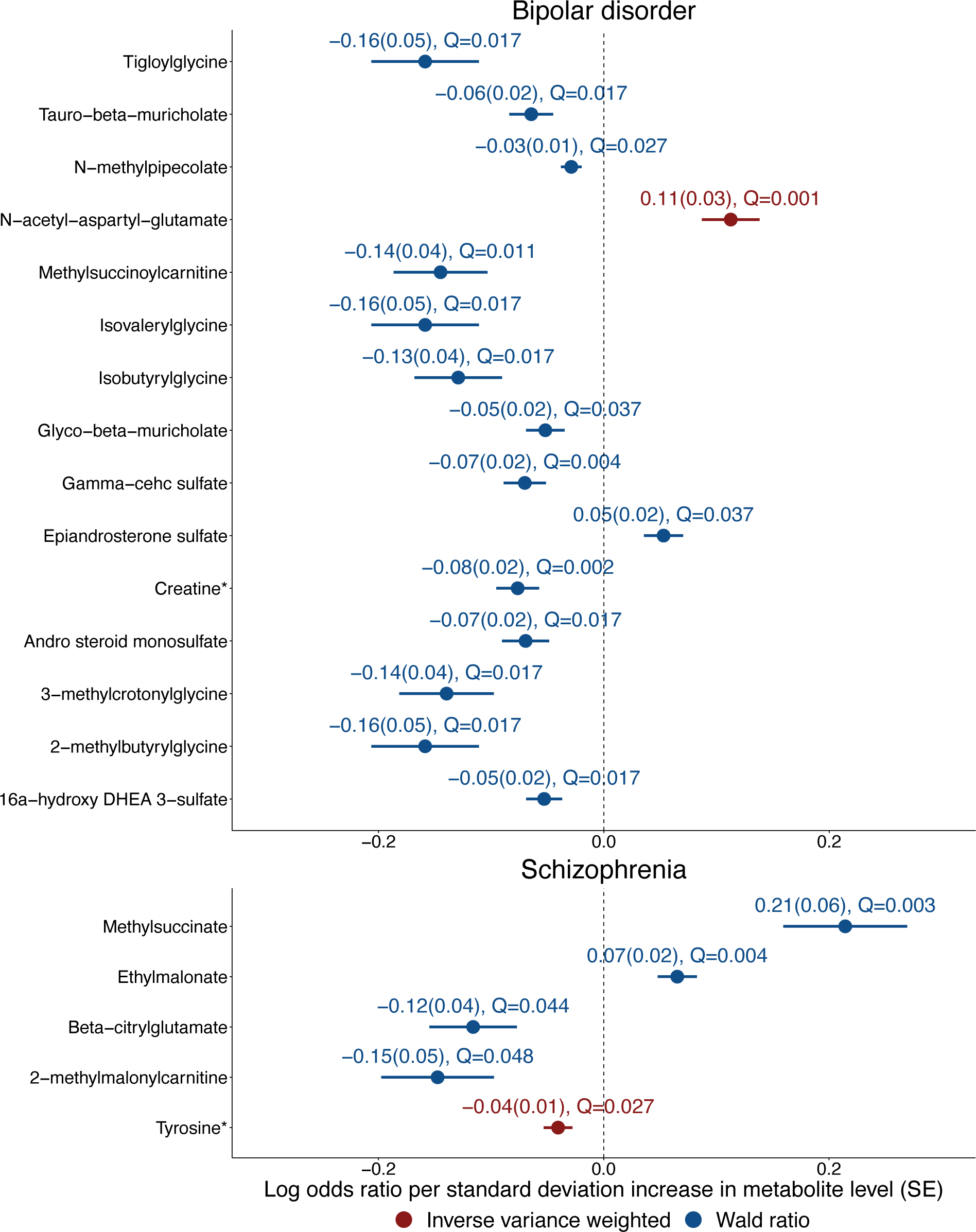
Estimated causal effects of urinary markers on the risk of bipolar disorder and schizophrenia. Urinary markers are shown on the Y-axis and log-odds ratio of bipolar disorder and schizophrenia are shown on the X-axis. Dots represent the mean effect size, with horizontal lines representing standard errors. Combined results from multiple studies are marked with an asterisk (*). Q-values represent P-values adjusted for multiple comparisons.

## 4. Discussion

The present study aimed to discover putative urinary metabolite biomarkers of psychiatric diseases using genetic instruments. To this end, a two-sample MR analysis was conducted using GWAS exposure data obtained from a meta-analysis of urinary metabolite GWAS studies as well as the publicly available GWAS catalog, and the outcome data obtained from the Psychiatric Genomics Consortium. Causal effects were identified for 67 analyte-disorder associations, of which 21 were specific to a single disease. Most notable findings of the analysis include the common association of N-acetylated compounds to BD and schizophrenia, tyrosine to schizophrenia, creatine to BD, N,N-dimethylglycine to ADHD, as well as pyridoxal and ferulic acid 4-sulfate to anorexia. The overlapping markers, as well as markers of ADHD and anorexia are discussed in the supplementary material in the Extended Discussion. Previously published direct and indirect associations were found in the existing literature for all of the shared putative biomarkers of the disorders. Furthermore, tyrosine and creatine have been reported to show strong and robust pathophysiological associations with schizophrenia and BD, respectively. Owing to their specificity and the multiple lines of supporting evidence, these biomarkers have potential to differentiate not only patients from healthy controls, but also different diseases from each other.

### 4.1. Specific biomarkers of bipolar disorder

The 15 significant urinary biomarkers associated uniquely with BD can be grouped into glycine-related compounds (creatine, tigloyl glycine, isovaleryl glycine, isobutyryl glycine, 3-methyl crotonyl glycine, and 2-methylbutyrylglycine), sulfates (gamma-cehc sulfate, epiandrosterone sulfate, 16A-hydroxy DHEA 3-sulfate, andro-steroid monosulfate), cholates (tauro-beta-muricholate, glyco-beta-muricholate), as well as individual compounds, N-acetyl-aspartyl-glutamate, N-methylpipecolate, and methyl succinyl carnitine. Although sulfates, cholates, and the individual compounds N-methylpipecolate and methyl succinyl carnitine have no known associations to BD, existing literature shows that glycine- and glutamate-related compounds have well-established links to the disease.

Glycine and glutamate alterations in plasma have been observed in BD patients during manic phases, irrespective of mood stabilizer treatment^45^. Glutamate and glycine are co-agonists at the N-methyl-D-aspartate receptor (NMDAR), the abnormalities of which have been hypothesized to be associated with symptoms of BD^46^. The specific details of the mechanism of action regarding NMDARs and BD are not yet thoroughly understood, however the results of the MR analysis further support the connection and suggest a pathophysiological role for N-acetyl-aspartyl-glutamate. Additionally, the MR analysis results imply that creatine is inversely correlated with BD. Creatine is a compound commonly linked with energy metabolism in muscle and brain tissue, and can be supplemented dietarily, however in the body it is also synthesized using glycine and other amino acids^47^. Creatine has been extensively linked with BD in past studies because of its relation to brain energy metabolism. Reductions of creatine levels have been identified in several regions of the brain in BD^48,49^, and total creatinine has been shown to be inversely correlated with depressive symptom severity in patients with BD^50^. In a clinical trial of creatine add-on supplementation, the efficacy of creatine in treating depressive symptoms was not statistically significant, however it was shown to significantly improve remission rates in BD patients^51^. Therefore, it can be hypothesized that naturally increased creatine levels could have protective effects in relation to symptoms of BD.

### 4.2. Specific biomarkers of schizophrenia

The five exclusive associations between urinary analytes and schizophrenia were tyrosine, methylsuccinate, ethylmalonate, beta-citrylglutamate, and 2-methylmalonylcarnitine. The analytes can be grouped into two categories. Tyrosine and beta-citrylglutamate are metabolically associated with the neurotransmitters dopamine and glutamate, which are central to the leading hypotheses of the mechanism of schizophrenia. In turn, 2-methylmalonylcarnitine, methylsuccinate, and ethylmalonate are associated with metabolic disturbances in mitochondria, which similarly have been hypothesised to cause schizophrenia.

Methyl succinic acid and ethylmalonic acid are conjugate acids of methylsuccinate and ethylmalonate, respectively. When upregulated in urine, they are biomarkers of the severe neurometabolic condition ethylmalonic encephalopathy (EE)^52^, which is characterized by disturbances in the brain, cardiovascular, and digestive systems. EE is caused by disturbances in mitochondrial function through mutations affecting the production of an enzyme which breaks down sulfides in mitochondria. As the MR results imply that upregulated urinary methylsuccinate and ethylmalonate increase susceptibility to schizophrenia, it can be hypothesized that a similar mechanism to that of EE involving mitochondrial dysfunction may exist in schizophrenia. In addition, ethylmalonate in cerebrospinal fluid has been directly associated with schizophrenia in the past^53^, suggesting that a mechanistic association may exist between ethylmalonate and the disease. In turn, 2-methylmalonylcarnitine is a short-chain acylcarnitine which has roles in acyl-group compound transport into mitochondria for energy production^54^, and it has been linked with ASC through effects on mitochondrial function^55^. Mitochondrial disturbances have been strongly linked with increased risk of schizophrenia^56,57^, however the exact pathomechanism remains elusive. The results of the MR analysis linking ethylmalonate, methylsuccinate, and 2-methylmalonylcarnitine to schizophrenia therefore indicate a possible causal linking factor through effects on mitochondrial function and should be further validated in future studies.

The MR results regarding tyrosine and beta-citrylglutamate are in line with the dopamine and glutamate hypotheses of schizophrenia, which are at present the dominant theories of the mechanisms underlying the disease^58,59^. Briefly, the dopamine hypothesis states that the psychotic symptoms of schizophrenia stem from hyperactive dopamine neurotransmission, as well as oversensitivity of dopamine receptors^59^. Most current antipsychotic treatments target this system through dopamine receptor blockage^59^. Tyrosine is a precursor amino acid of dopamine and is thus central to the dopaminergic pathway, acting as a limiting factor in the production of dopamine^60^. Observed causal effects from the MR imply that higher levels of tyrosine result in a lower likelihood of schizophrenia development. The results can be interpreted within the context of the dopamine hypothesis as being a result of potentially enhanced conversion of tyrosine into dopamine, however other possibilities also exist. Previous clinical trials have not supported the possibility of a protective effect of tyrosine on schizophrenia symptoms^61^, therefore it is more likely that the observed effect is an outcome of a mechanistic step in the symptom development pathway. Other studies have suggested that limited tyrosine transport to the brain through the blood brain barrier could be causal to the alterations of dopamine levels in the brains of schizophrenia patients^62,63^. Therefore, increased peripheral levels of tyrosine could imply that an excess of tyrosine may mitigate the lower tyrosine transport into the brain, and thus reduce the symptoms of schizophrenia.

The literature regarding beta-citrylglutamate is limited, however it is a known derivative of glutamate, and concentrations of beta-citrylglutamate decrease with age^64^. The glutamate hypothesis of schizophrenia is similar to the observations of glutamate disruptions in BD, where the downregulated glutamate signalling function of NDMARs are hypothesized to be associated with the cognitive and negative symptoms of schizophrenia^58^. The MR results imply that decreased urinary beta-citrylglutamate increases the risk of developing schizophrenia, which may be due to lower baseline levels of glutamate resulting in decreased glutamate signalling. Taken together, the present results are in line with the dopamine and glutamate hypotheses of schizophrenia and indicate urinary tyrosine and glutamate as potential biomarkers.

### 4.3. Urinary diagnostics

The observed urinary analyte-disorder associations, if validated, have the potential to be used in a differential diagnostic biomarker panel. This is due to the sheer number of markers exclusive to a single disease, specifically for BD and schizophrenia. The collective predictive value of the identified biomarkers could surpass the threshold for utility in a clinical test. A significant benefit of the discovered biomarkers concerns the target disorders. BD and schizophrenia are commonly misdiagnosed with each other, with an estimated 24% of schizophrenia patients being misdiagnosed with BD^12,65^, and one-third of BD patients receiving an initial diagnosis of schizophrenia^66^. Therefore, a diagnostic panel would only need to be able to accurately differentiate the two disorders from each other to be clinically relevant. Additionally, for high-risk individuals such as close relatives of patients with the respective disorders, a point-of-care diagnostic assessment could be developed to identify early indicators of either disorder, owing to the non-invasive nature of urinary measurements.

### 4.4. Computational screening for biomarker discovery

The present study highlights the robust capability of computational methods to pre-emptively identify high-priority targets in biomarker discovery in addition to traditional methods. The current landscape in biomarker discovery does not utilize computational resources to its full extent. While MR studies have been carried out in the past for biomarkers of mental health disorders in the past^67,68^, the intent is often to exclusively identify causal effects and not disease biomarkers. Additionally, in the vast majority of biomarker discovery trials, computational screening is not utilized for target identification. As the library of GWAS studies expands, the use of computational biological assessment through MR should be more accessible and applicable to a wide variety of disorders and markers. Therefore, the use of computational methods for biomarker candidate screening should be broadly implemented to maximize the probability of success in clinical trials, especially considering the high costs of conducting such studies, and the low cost of MR analysis.

### 4.5. Limitations

Urinary analyte levels are significantly impacted both between and within individuals and depend on changes in daily activities such as diet, hydration, or activity levels^16,17^. When considering that the majority of the identified markers show small effect sizes, the observation and validation of the markers in real samples could be challenging. Another limitation is the lack of availability of comprehensive urinary GWAS data. Urinary analyte GWAS compared to other GWAS studies typically have small sample sizes, measure a limited number of analytes, and use more variable scales, which limits the number of SNPs available for analysis and necessitates the use of non-standard MR methods, such as the one used in the present study. Additionally, palindromicity of the analysed SNPs was not evaluated in the analysis due to a lack of allele frequency data in the majority of the exposure datasets. In the outcome datasets, the SNP associations are assessed against healthy controls and not against other disorder groups, therefore it is feasible that some differential biomarkers identified in this study may overlap in real patient samples. Finally, the assessed GWAS studies in the analysis are primarily based on European populations, therefore the analysis might not translate to other ethnic populations.

### 4.6. Conclusions

In conclusion, we identified 21 urinary biomarker candidates with a potential for differential diagnosis of psychiatric disorders using a two-sample MR approach. Of the putative biomarkers, five were associated with schizophrenia, 15 with BD, and one with ADHD. Tyrosine for schizophrenia and creatine for BD were considered the most robust markers, owing to their effects being identified based on multiple studies. The findings indicate that urine could be a valuable source of biomarkers for various psychiatric disorders and highlight the strength of computational and genetic analyses for the purposes of biomarker discovery. With further validation, the identified markers could be used for developing a differential diagnostic non-invasive biomarker platform for psychiatric diseases, which is much needed to lower delays in diagnosis as well as misdiagnosis rates of severe psychiatric disorders and mitigate the significant economic and individual burdens.

## Data Availability

All data produced in this study is available from the corresponding authors upon reasonable request.

## References

1 Ferrari A, Santomauro D, Mantilla Herrera A, et al. Global, regional, and national burden of 12 mental disorders in 204 countries and territories, 1990–2019: a systematic analysis for the Global Burden of Disease Study 2019. Lancet Psychiatry 2022; 9: 137–50.

2 Department of Health. Annual Report of the Chief Medical Officer 2013 Public Mental Health Priorities: Investing in the Evidence. 2014. gov.uk/government/publications/chief-medical-officer-cmoannual-report-public-mental-health (accessed Nov 25, 2022).

3 Mccrone P, Dhanasiri S, Patel A, Knapp M, Lawton-Smith S. Paying the Price: the cost of mental health care in England in 2026. The Kings Fund 2008; : 15–34.

4 Simon J, Pari AAA, Wolstenholme J, Berger M, Goodwin GM, Geddes JR. The costs of bipolar disorder in the United Kingdom. Brain Behav 2021; 11: e2351.

5 Mangalore R, Knapp M. Cost of schizophrenia in England. J Ment Health Policy Econ 2007; 10: 23–41.

6 Hung CI, Liu CY, Yang CH. Untreated duration predicted the severity of depression at the two-year follow-up point. PLoS One 2017; 12. DOI:10.1371/JOURNAL.PONE.0185119.

7 Vieta E, Salagre E, Grande I, et al. Early Intervention in Bipolar Disorder. Am J Psychiatry 2018; 175: 411–26.

8 Lieberman JA, Small SA, Girgis RR. Early Detection and Preventive Intervention in Schizophrenia: From Fantasy to Reality. American Journal of Psychiatry 2019; 176: 794–810.

9 Insel T, Cuthbert B, Garvey M, et al. Research Domain Criteria (RDoC): Toward a New Classification Framework for Research on Mental Disorders. 10.1176/appi.ajp201009091379 2010; 167: 748–51.

10 Cheniaux E, Landeira-Fernandez J, Versiani M. The Diagnoses of Schizophrenia, Schizoaffective Disorder, Bipolar Disorder and Unipolar Depression: Interrater Reliability and Congruence between DSM-IV and ICD-10. Psychopathology 2009; 42: 293–8.

11 Feczko E, Miranda-Dominguez O, Marr M, Graham AM, Nigg JT, Fair DA. The Heterogeneity problem: Approaches to identify psychiatric subtypes. Trends Cogn Sci 2019; 23: 584–601.

12 Ayano G, Demelash S, yohannes Z, et al. Misdiagnosis, detection rate, and associated factors of severe psychiatric disorders in specialized psychiatry centers in Ethiopia. Ann Gen Psychiatry 2021; 20: 1–10.

13 Lubloy Á, Keresztúri JL, Ńemeth A, Mihalicza P. Exploring factors of diagnostic delay for patients with bipolar disorder: A population-based cohort study. BMC Psychiatry 2020; 20: 1–17.

14 Norman RMG, Malla AK, Verdi MB, Hassall LD, Fazekas C. Understanding delay in treatment for first-episode psychosis. Psychol Med 2004; 34: 255–66.

15 Harpole M, Davis J, Espina V. Current state of the art for enhancing urine biomarker discovery. 10.1080/1478945020161190651 2016; 13: 609–26.

16 Chen Y. Variations of human urinary proteome. Adv Exp Med Biol 2015; 845: 91–4.

17 Nagaraj N, Mann M. Quantitative analysis of the intra- and inter-individual variability of the normal urinary proteome. J Proteome Res 2011; 10: 637–45.

18 Davies NM, Holmes M v., Davey Smith G. Reading Mendelian randomisation studies: a guide, glossary, and checklist for clinicians. BMJ 2018; 362: 601.

19 Zaki J, Tomasik J, Scherman O, Bahn S. Meta-Analysis of Urinary Metabolite GWAS Identifies Two Novel Genome-Wide Significant Loci. Manuscript in preparation.

20 Raffler J, Friedrich N, Arnold M, et al. Genome-Wide Association Study with Targeted and Non-targeted NMR Metabolomics Identifies 15 Novel Loci of Urinary Human Metabolic Individuality. PLoS Genet 2015; 11: e1005487.

21 Schlosser P, Li Y, Sekula P, et al. Genetic studies of urinary metabolites illuminate mechanisms of detoxification and excretion in humans. Nat Genet 2020; 52: 167–76.

22 Nicholson G, Rantalainen M, Li J v., et al. A Genome-Wide Metabolic QTL Analysis in Europeans Implicates Two Loci Shaped by Recent Positive Selection. PLoS Genet 2011; 7: e1002270.

23 Calvo-Serra B, Maitre L, Lau CHE, et al. Urinary metabolite quantitative trait loci in children and their interaction with dietary factors. Hum Mol Genet 2021; 29: 3830–44.

24 Sinnott-Armstrong N, Tanigawa Y, Amar D, et al. Genetics of 35 blood and urine biomarkers in the UK Biobank. Nat Genet 2021; 53: 185–94.

25 Willer CJ, Li Y, Abecasis GR. METAL: fast and efficient meta-analysis of genomewide association scans. Bioinformatics 2010; 26: 2190–1.

26 Li Y, Sekula P, Wuttke M, et al. Genome-Wide Association Studies of Metabolites in Patients with CKD Identify Multiple Loci and Illuminate Tubular Transport Mechanisms. Journal of the American Society of Nephrology 2018; 29: 1513–24.

27 Rueedi R, Ledda M, Nicholls AW, et al. Genome-Wide Association Study of Metabolic Traits Reveals Novel Gene-Metabolite-Disease Links. PLoS Genet 2014; 10: e1004132.

28 Sekula P, Tin A, Schultheiss UT, et al. Urine 6-Bromotryptophan: Associations with Genetic Variants and Incident End-Stage Kidney Disease. Scientific Reports 2020 10:1 2020; 10: 1–11.

29 Comuzzie AG, Cole SA, Laston SL, et al. Novel Genetic Loci Identified for the Pathophysiology of Childhood Obesity in the Hispanic Population. PLoS One 2012; 7: e51954.

30 Olden M, Corre T, Hayward C, et al. Common variants in UMOD associate with urinary uromodulin levels: A meta-analysis. Journal of the American Society of Nephrology 2014; 25: 1869–82.

31 Demontis D, Walters RK, Martin J, et al. Discovery of the first genome-wide significant risk loci for attention deficit/hyperactivity disorder. Nat Genet 2018; 51: 63–75.

32 Watson HJ, Yilmaz Z, Thornton LM, et al. Genome-wide association study identifies eight risk loci and implicates metabo-psychiatric origins for anorexia nervosa. Nat Genet 2019; 51: 1207–14.

33 Grove J, Ripke S, Als TD, et al. Identification of common genetic risk variants for autism spectrum disorder. Nat Genet 2019; 51: 431–44.

34 Mullins N, Forstner AJ, O’Connell KS, et al. Genome-wide association study of more than 40,000 bipolar disorder cases provides new insights into the underlying biology. Nat Genet 2021; 53: 817–29.

35 Giannakopoulou O, Lin K, Meng X, et al. The Genetic Architecture of Depression in Individuals of East Asian Ancestry: A Genome-Wide Association Study. JAMA Psychiatry 2021; 78: 1258–69.

36 Trubetskoy V, Pardiñas AF, Qi T, et al. Mapping genomic loci implicates genes and synaptic biology in schizophrenia. Nature 2022; 604: 502–8.

37 Yu D, Sul JH, Tsetsos F, et al. Interrogating the genetic determinants of Tourette’s syndrome and other tiC disorders through genome-wide association studies. American Journal of Psychiatry 2019; 176: 217–27.

38 Hemani G, Zheng J, Elsworth B, et al. The MR-base platform supports systematic causal inference across the human phenome. Elife 2018; 7: e34408.

39 Auton A, Abecasis GR, Altshuler DM, et al. A global reference for human genetic variation. Nature 2015; 526: 68–74.

40 Machiela MJ, Chanock SJ. LDlink: a web-based application for exploring population-specific haplotype structure and linking correlated alleles of possible functional variants. Bioinformatics 2015; 31: 3557.

41 Sanderson E, Glymour MM, Holmes M v., et al. Mendelian randomization. Nature Reviews Methods Primers 2022; 2: 6.

42 Bowden J, Davey Smith G, Haycock PC, Burgess S. Consistent Estimation in Mendelian Randomization with Some Invalid Instruments Using a Weighted Median Estimator. Genet Epidemiol 2016; 40: 304–14.

43 Kamat MA, Blackshaw JA, Young R, et al. PhenoScanner V2: an expanded tool for searching human genotype–phenotype associations. Bioinformatics 2019; 35: 4851–3.

44 Staley JR, Blackshaw J, Kamat MA, et al. PhenoScanner: a database of human genotype–phenotype associations. Bioinformatics 2016; 32: 3207–9.

45 Hoekstra R, Fekkes D, Loonen AJM, Pepplinkhuizen L, Tuinier S, Verhoeven WMA. Bipolar mania and plasma amino acids: increased levels of glycine. Eur Neuropsychopharmacol 2006; 16: 71–7.

46 Ghasemi M, Phillips C, Trillo L, de Miguel Z, Das D, Salehi A. The role of NMDA receptors in the pathophysiology and treatment of mood disorders. Neurosci Biobehav Rev 2014; 47: 336–58.

47 Wyss M, Kaddurah-Daouk R. Creatine and creatinine metabolism. Physiol Rev 2000; 80: 1107–213.

48 Kious BM, Kondo DG, Renshaw PF. Creatine for the Treatment of Depression. Biomolecules 2019, Vol 9, Page 406 2019; 9: 406.

49 Allen PJ. Creatine metabolism and psychiatric disorders: Does creatine supplementation have therapeutic value? Neurosci Biobehav Rev 2012; 36: 1442–62.

50 Dager SR, Friedman SD, Parow A, et al. Brain Metabolic Alterations in Medication-Free Patients With BipolarDisorder. Arch Gen Psychiatry 2004; 61: 450–8.

51 Toniolo RA, Silva M, Fernandes F de BF, Amaral JA de MS, Dias R da S, Lafer B. A randomized, double-blind, placebo-controlled, proof-of-concept trial of creatine monohydrate as adjunctive treatment for bipolar depression. J Neural Transm 2018; 125: 247–57.

52 Drousiotou A, DiMeo I, Mineri R, Georgiou T, Stylianidou G, Tiranti V. Ethylmalonic encephalopathy: application of improved biochemical and molecular diagnostic approaches. Clin Genet 2011; 79: 385–90.

53 Panyard DJ, Kim KM, Darst BF, et al. Cerebrospinal fluid metabolomics identifies 19 brain-related phenotype associations. Commun Biol 2021; 4: 1–11.

54 Indiveri C, Iacobazzi V, Tonazzi A, et al. The mitochondrial carnitine/acylcarnitine carrier: Function, structure and physiopathology. Mol Aspects Med 2011; 32: 223–33.

55 Kepka A, Ochocinska A, Chojnowska S, et al. Potential Role of L-Carnitine in Autism Spectrum Disorder. J Clin Med 2021; 10: 1–26.

56 Flippo KH, Strack S. An emerging role for mitochondrial dynamics in schizophrenia. Schizophr Res 2017; 187: 26–32.

57 Prabakaran S, Swatton JE, Ryan MM, et al. Mitochondrial dysfunction in schizophrenia: evidence for compromised brain metabolism and oxidative stress. Mol Psychiatry 2004; 9: 684–97.

58 Coyle JT. The glutamatergic dysfunction hypothesis for schizophrenia. Harv Rev Psychiatry 1996; 3: 241–53.

59 Howes OD, Kapur S. The Dopamine Hypothesis of Schizophrenia: Version III—The Final Common Pathway. Schizophr Bull 2009; 35: 549–62.

60 Daubner SC, Le T, Wang S. Tyrosine Hydroxylase and Regulation of Dopamine Synthesis. Arch Biochem Biophys 2011; 508: 12.

61 Deutsch SI, Rosse RB, Schwartz BL, Banay-Schwartz M, McCarthy MF, Johri SK. L-tyrosine pharmacotherapy of schizophrenia: preliminary data. Clin Neuropharmacol 1994; 17: 53–62.

62 Wiesel FA, Andersson JLR, Westerberg G, et al. Tyrosine transport is regulated differently in patients with schizophrenia. Schizophr Res 1999; 40: 37–42.

63 Bjerkenstedt L, Farde L, Terenius L, Edman G, Venizelos N, Wiesel FA. Support for limited brain availability of tyrosine in patients with schizophrenia. Int J Neuropsychopharmacol 2006; 9: 247–55.

64 Collard F, Stroobant V, Lamosa P, et al. Molecular Identification of N-Acetylaspartylglutamate Synthase and *β*-Citrylglutamate Synthase. Journal of Biological Chemistry 2010; 285: 29826–33.

65 Lopez-Castroman J, Leiva-Murillo JM, Cegla-Schvartzman F, et al. Onset of schizophrenia diagnoses in a large clinical cohort. Sci Rep 2019; 9: 9865.

66 Gonzalez-Pinto A, Gutierrez M, Mosquera F, et al. First episode in bipolar disorder: misdiagnosis and psychotic symptoms. J Affect Disord 1998; 50: 41–4.

67 Yang J, Yan B, Zhao B, et al. Assessing the Causal Effects of Human Serum Metabolites on 5 Major Psychiatric Disorders. Schizophr Bull 2020; 46: 804–13.

68 Lu T, Forgetta V, Greenwood CMT, Zhou S, Richards JB. Circulating Proteins Influencing Psychiatric Disease: A Mendelian Randomization Study. Biol Psychiatry 2023; 93: 82–91.

