## Supplementary material for "Discovery of Urinary Metabolite Biomarkers of Psychiatric Disorders Using Two-Sample Mendelian Randomization"

**Supplementary Table 1. Results of the Mendelian randomization analysis for all significant causal estimates.** WR = Wald ratio method, IVW = inverse variance weighted ratio method, N = number of studies assessed, Q = Benjamini-Hochberg adjusted significance threshold.

| Exposure | Outcome | Method | Beta | SE | Q | N |
|---|---|---|---|---|---|---|
| HOMOCITRULLINE | SCZ | WR | -0.12 | 0.02 | 3.56E-05 | 1 |
| N-ACETYL-3-METHYLHISTIDINE | SCZ | WR | 0.06 | 0.01 | 3.56E-05 | 1 |
| N-ACETYLASPARAGINE | SCZ | WR | 0.06 | 0.01 | 3.56E-05 | 1 |
| N-ACETYLGLUTAMINE | SCZ | WR | 0.06 | 0.01 | 3.56E-05 | 1 |
| N-ACETYLHISTIDINE | SCZ | WR | 0.22 | 0.04 | 3.56E-05 | 1 |
| N-ACETYLKYNURENINE 2 | SCZ | WR | 0.06 | 0.01 | 3.56E-05 | 1 |
| N-ACETYLLEUCINE | SCZ | WR | 0.11 | 0.02 | 3.56E-05 | 1 |
| N-ACETYLPYRRALINE | SCZ | WR | 0.07 | 0.01 | 3.56E-05 | 1 |
| N-ALPHA-ACETYLORNITHINE | SCZ | WR | 0.14 | 0.03 | 3.56E-05 | 1 |
| N-DELTA-ACETYLORNITHINE | SCZ | WR | -0.05 | 0.01 | 3.56E-05 | 1 |
| N2-ACETYLLYSINE | SCZ | WR | 0.06 | 0.01 | 3.56E-05 | 1 |
| N2,N6-DIACETYLLYSINE | SCZ | WR | 0.07 | 0.01 | 3.56E-05 | 1 |
| N-ACETYL-1-METHYLHISTIDINE | SCZ | WR | 0.07 | 0.01 | 4.29E-05 | 1 |
| N-ACETYLARGININE | SCZ | WR | 0.09 | 0.02 | 4.29E-05 | 1 |
| N-ACETYLCITRULLINE | SCZ | WR | 0.05 | 0.01 | 4.29E-05 | 1 |
| PICOLINATE | SCZ | WR | 0.16 | 0.03 | 5.20E-05 | 1 |
| CYSTEINYLGLYCINE DISULFIDE | SCZ | WR | -0.19 | 0.04 | 5.86E-05 | 1 |
| N6-CARBOXYETHYLLYSINE | SCZ | WR | -0.25 | 0.05 | 5.86E-05 | 1 |
| PROLYLGLYCINE | SCZ | WR | -0.05 | 0.01 | 5.86E-05 | 1 |
| ADENOSINE 3',5'-CYCLIC MONOPHOSPHATE | SCZ | WR | 0.19 | 0.04 | 8.94E-05 | 1 |
| N-ACETYL-ASPARTYL-GLUTAMATE | BD | IVW | 0.11 | 0.03 | 0.001 | 1 |
| N-ACETYLHISTIDINE | BD | WR | 0.22 | 0.05 | 0.001 | 1 |
| N-ACETYLKYNURENINE 2 | BD | WR | 0.06 | 0.01 | 0.001 | 1 |
| CREATINE | BD | WR | -0.08 | 0.02 | 0.002 | 3 |
| PYRIDOXAL | BD | WR | -0.09 | 0.02 | 0.003 | 1 |
| FERULIC ACID 4-SULFATE | BD | WR | -0.07 | 0.02 | 0.003 | 1 |
| HOMOCITRULLINE | BD | WR | -0.11 | 0.03 | 0.003 | 1 |
| METHYLSUCCINATE | SCZ | WR | 0.21 | 0.06 | 0.003 | 1 |
| N-ACETYL-3-METHYLHISTIDINE | BD | WR | 0.05 | 0.01 | 0.003 | 1 |
| N-ACETYLASPARAGINE | BD | WR | 0.06 | 0.01 | 0.003 | 1 |
| N-ACETYLGLUTAMINE | BD | WR | 0.05 | 0.01 | 0.003 | 1 |
| N-ACETYLLEUCINE | BD | WR | 0.10 | 0.03 | 0.003 | 1 |
| N-ACETYLPYRRALINE | BD | WR | 0.06 | 0.01 | 0.003 | 1 |
| N-ALPHA-ACETYLORNITHINE | BD | WR | 0.12 | 0.03 | 0.003 | 1 |
| N-DELTA-ACETYLORNITHINE | BD | WR | -0.04 | 0.01 | 0.003 | 1 |
| N2-ACETYLLYSINE | BD | WR | 0.05 | 0.01 | 0.003 | 1 |
| N2,N6-DIACETYLLYSINE | BD | WR | 0.06 | 0.02 | 0.003 | 1 |
| ADENOSINE 3',5'-CYCLIC MONOPHOSPHATE | BD | WR | 0.19 | 0.05 | 0.003 | 1 |
| N6-CARBOXYETHYLLYSINE | BD | WR | -0.22 | 0.06 | 0.004 | 1 |
| N-ACETYL-1-METHYLHISTIDINE | BD | WR | 0.06 | 0.02 | 0.004 | 1 |
| N-ACETYLARGININE | BD | WR | 0.08 | 0.02 | 0.004 | 1 |
| N-ACETYLCITRULLINE | BD | WR | 0.05 | 0.01 | 0.004 | 1 |

| | | | | | | |
|---|---|---|---|---|---|---|
| ETHYLMALONATE | SCZ | WR | 0.07 | 0.02 | 0.004 | 1 |
| GAMMA-CEHC SULFATE | BD | WR | -0.07 | 0.02 | 0.004 | 1 |
| N,N-DIMETHYLGLYCINE | ADHD | WR | -0.39 | 0.11 | 0.008 | 1 |
| METHYLSUCCINOYLCARNITINE | BD | WR | -0.14 | 0.04 | 0.011 | 1 |
| 16A-HYDROXY DHEA 3-SULFATE | BD | WR | -0.05 | 0.02 | 0.017 | 1 |
| 2-METHYLBUTYRYLGLYCINE | BD | WR | -0.16 | 0.05 | 0.017 | 1 |
| 3-METHYLCROTONYLGLYCINE | BD | WR | -0.14 | 0.04 | 0.017 | 1 |
| ANDRO STEROID MONOSULFATE | BD | WR | -0.07 | 0.02 | 0.017 | 1 |
| ISOBUTYRYLGLYCINE | BD | WR | -0.13 | 0.04 | 0.017 | 1 |
| ISOVALERYLGLYCINE | BD | WR | -0.16 | 0.05 | 0.017 | 1 |
| TAURO-BETA-MURICHOLATE | BD | WR | -0.06 | 0.02 | 0.017 | 1 |
| TIGLOYLGLYCINE | BD | WR | -0.16 | 0.05 | 0.017 | 1 |
| FERULIC ACID 4-SULFATE | SCZ | WR | -0.05 | 0.01 | 0.020 | 1 |
| PYRIDOXAL | SCZ | WR | -0.07 | 0.02 | 0.022 | 1 |
| TYROSINE | SCZ | IVW | -0.04 | 0.01 | 0.027 | 3 |
| N-METHYLPIPECOLATE | BD | WR | -0.03 | 0.01 | 0.027 | 1 |
| PICOLINATE | BD | WR | 0.13 | 0.04 | 0.027 | 1 |
| FERULIC ACID 4-SULFATE | ANO | WR | 0.08 | 0.03 | 0.037 | 1 |
| CYSTEINYLGLYCINE DISULFIDE | BD | WR | -0.15 | 0.05 | 0.037 | 1 |
| EPIANDROSTERONE SULFATE | BD | WR | 0.05 | 0.02 | 0.037 | 1 |
| GLYCO-BETA-MURICHOLATE | BD | WR | -0.05 | 0.02 | 0.037 | 1 |
| PROLYLGLYCINE | BD | WR | -0.04 | 0.01 | 0.037 | 1 |
| PYRIDOXAL | ANO | WR | 0.10 | 0.03 | 0.042 | 1 |
| BETA-CITRYLGLUTAMATE | SCZ | WR | -0.12 | 0.04 | 0.044 | 1 |
| 2-METHYLMALONYLCARNITINE | SCZ | WR | -0.15 | 0.05 | 0.048 | 1 |

**Supplementary Table 2. Heterogeneity statistics for the associations of tyrosine to schizophrenia and N-acetyl-aspartyl-glutamate to bipolar disorder.** Q = Cochran's Q statistic, IVW = inverse variance weighted method.

| Exposure | Disorder | Method | Q | P-value |
|---|---|---|---|---|
| TYROSINE | SCZ | IVW | 1.75 | 0.19 |
| TYROSINE | SCZ | IVW | 1.78 | 0.41 |
| TYROSINE | SCZ | IVW | 0.50 | 0.78 |
| N-ACETYL-ASPARTYL-GLUTAMATE | BD | IVW | 0.00 | 0.99 |

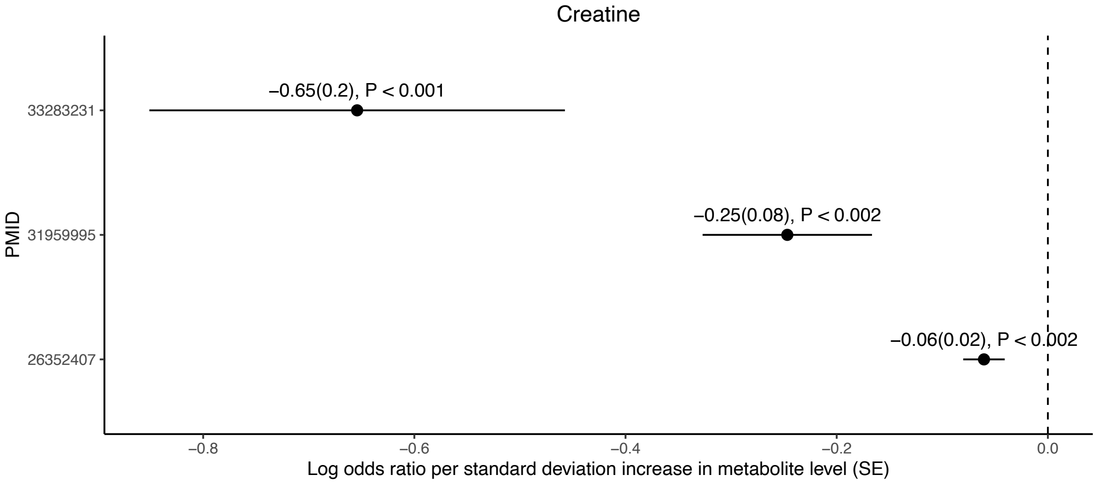


**Supplementary Figure 1. Individual study results for the association of creatine to bipolar disorder.** Dots represent the beta coefficient, and the lines represent standard errors. Significance levels have been shown as P-values.

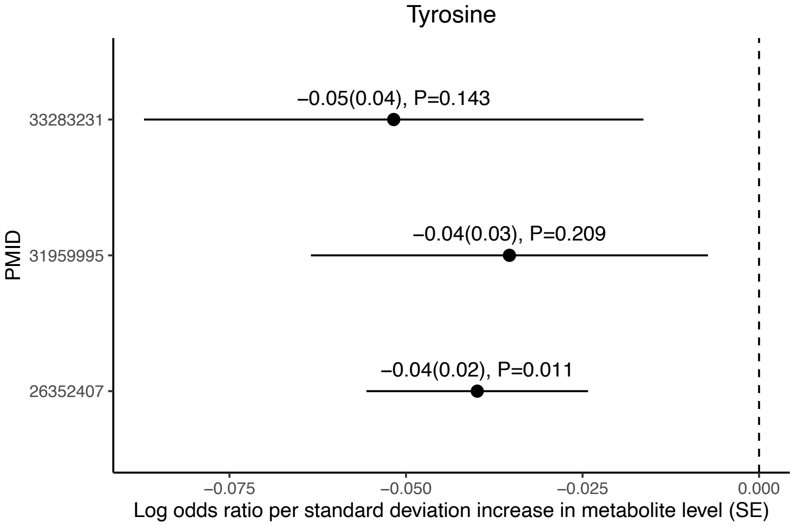


**Supplementary Figure 2. Individual study results for the association of tyrosine to schizophrenia.** Dots represent the beta coefficient, and the lines represent standard errors. Significance levels have been shown as P-values.

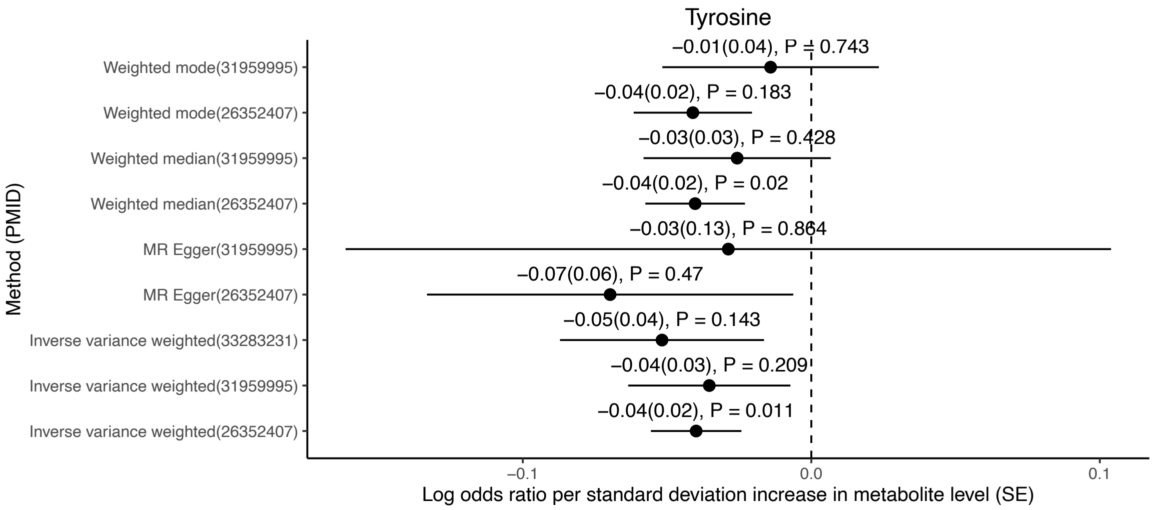


**Supplementary Figure 3. Effect size consistency analysis for the association of tyrosine to schizophrenia.** Dots represent the beta coefficient, and the lines represent standard errors. Significance levels have been shown as P-values.

### Extended Discussion

#### *1.1 Markers of ADHD and anorexia nervosa*

N,N-dimethylglycine, or DMG, is a glycine derivative produced within the body, and is commonly marketed as a dietary supplement for a wide range of indications including ADHD and ASC, however scientific literature supporting its use is limited. Few clinical trials of DMG have been conducted for various purposes such as the treatment of symptoms of multiple sclerosis[1] and ASC[2], however limited improvements in cognitive symptoms have been observed. While past literature is scarce regarding the supplementation of DMG for the treatment of ADHD, the MR results imply that a decrease in urinary DMG increases the risk of ADHD. This in turn implies a putative therapeutic or protective effect of DMG on symptoms of ADHD.

Pyridoxal, also known as vitamin $B_6$, is a critical compound with important roles in the biosynthesis of amino acids, fatty acids, as well as the breakdown of carbohydrate storage compounds[3]. General vitamin B deficiencies have been identified as a risk factor in developing anorexia nervosa nearly a century ago[4], and in a more recent research study, 31% of assessed anorexia patients had indicators of vitamin $B_6$ deficiency[5]; however, the initial study was conducted in an animal model and the second study only had a sample size of 13 patients. Contrary to the above-mentioned studies, the MR results indicate that an increase in urinary vitamin

$B_6$ increases the risk of anorexia. One possibility is that the increased rate of vitamin $B_6$ excretion is a causal factor for the vitamin $B_6$ deficiency present in some forms of anorexia. Further investigations are necessary to understand the mechanisms of action in vitamin $B_6$-related anorexia nervosa.

In addition, the current study found that higher genetically predicted levels of ferulic acid 4-sulfate are associated with an increased risk of anorexia. Ferulic acid 4-sulfate is a metabolite of ferulic acid, which has been linked in the past with appetite control. Ferulic acid is commonly found in fruits and coffee, which are commonly consumed in attempts to control appetite, and past animal studies have identified a short-term 70% decrease in appetite when ferulic acid was injected[6]. Additionally, it has been observed that ferulic acid intake lowered body weight and fat accumulation in mice studies[7], which further support the association of ferulic acid with the development of anorexia. The associated effects on decreased appetite and weight gain, could further aggravate the core symptoms such as excessive loss of body weight. These results indicate potential molecular mediators and may provide valuable insights into the mechanistic underpinnings of anorexia.

#### *1.2 Shared markers of bipolar disorder and schizophrenia*

Schizophrenia and BD showed overlapping associations with 22 unique analytes, which can be categorized into n-acetylated amino acid related compounds (11), amino acid related compounds (picolinate, prolylglycine, cysteinylglycine disulfide, and homocitrulline), advanced glycation endproducts (AGEs) N6-carboxyethyllysine and N-acetylpyrraline, as well as the individual compounds ferulic acid 4-sulfate, pyridoxal, and cyclic adenosine monophosphate (cAMP). Past direct or indirect associations to both BD and schizophrenia have been reported for all of the non-acetylated compounds as well as multiple N-acetylated compounds, as described below.

The abundance of associations with N-acetylated compounds in the MR results indicate a potential link between the N-acetylation mechanism of amino acids and both schizophrenia and BD. The mechanism of free amino acid acetylation is not currently well understood; however they are most likely products of degraded acetylated proteins. The N-terminal acetylation of amino acids is an irreversible process, which alters the basic properties of proteins through the addition of an acetyl group to the N-terminal of an amino group catalysed by N-acetyltransferase (NAT) enzymes[8]. N-acetylation has been identified to be central in protein folding, half-life regulation, and complex formation, as well as membrane targeting and histone modification. Different N-acetylated compounds have been associated in the past literature with both schizophrenia[9,10] and BD[10,11], and polymorphisms in the N-acetyltransferase-2 (*NAT2*) gene which encodes for NATs have been associated with schizophrenia[12,13]. Due to the number of N-acetylated compounds associated with both BD and schizophrenia in the MR analysis, as well as the further association of schizophrenia to NAT encoding genes, it could be hypothesised that the links stem from dysfunctional NAT function instead of discrepancies in any N-acetylated compound alone.

Amino acid-related compounds have been associated with BD and schizophrenia numerous times in the existing literature[11,14,15]. Both of the non-acetylated glycine derivative compounds, prolylglycine and cycsteinylglycine disulfide, can be linked to BD and schizophrenia through NDMAR abnormalities, as mentioned in the discussion section. Additionally, a cyclic form of prolylglycine has been associated with antidepressant-like effects in animal models[16]. The results of the MR analysis indicate that upregulated expression of urinary prolylglycine could be protective of BD and schizophrenia, which may be a result of potentially reduced depressive symptoms in both disorders. Picolinate in turn is a conjugate base of picolinic acid, a metabolite of the amino acid tryptophan, which has been directly associated in the past with both schizophrenia and BD[17,18]. Tryptophan is a metabolic precursor to kynurenine, which acts as an antagonist at NDMARs[15] and is central to the kynurenine hypothesis of BD and schizophrenia[15]. In addition to picolinate, N-acetylkynurenine was significantly associated with both schizophrenia and BD in the MR analysis, further supporting the involvement of the kynurenine pathway and indicating a notable association with the urinary excretion of the tryptophan metabolic pathway products. Finally, homocitrulline is an amino acid metabolite of ornithine which has been directly associated with schizophrenia in plasma[19], and BD in serum[11]. Homocitrulline and ornithine are central to the citric and urea cycles, which when impaired have been associated with psychosis in general[20], and neurological symptoms in animal models[21].

AGEs dysregulation has been associated in the past with BD[22], and  accumulation of AGEs in body tissues and blood has been associated with schizophrenia in past literature[23]. AGE accumulation is an outcome of dietary intake of ultra-processed foods as well as aging, and their formation is driven by exposure to various proteins and lipids to glucose. While the MR results regarding N-acetylpyrraline are consistent with past findings regarding its accumulation, N6-carboxyethyllysine is shown to have a negative estimate, which would imply that a surplus of N6-carboxyethyllysine is preventative of both BD and schizophrenia. One possibility is that the increased urinary concentrations of N6-carboxyethyllysine implies an improved excretion rate, lowering its accumulation in the rest of the body, while the inverse estimate of N-acetylpyrraline could be an effect of the above-mentioned dysfunctional N-acetylation. While the association of AGEs to schizophrenia is robust, further research is recommended regarding their association with BD as well as the specific mechanisms underlying N6-carboxyethyllysine effects in both disorders.

Vitamin B has been hypothesized to be a potential supplement in the treatment of BD and schizophrenia, and vitamin $B_6$ as well as other forms of vitamin B have been identified to reduce psychiatric symptoms of schizophrenia in various clinical trials[24], however it has not been found effective as a supplement for treating BD symptoms[25]. Additionally, polymorphisms in vitamin B-related genes have been associated with both BD and schizophrenia, as well as other neuropsychiatric disorders[26]. Ferulic acid in turn, while not directly associated with either of the disorders, has been shown to have antidepressant-like effects in animal models through its antioxidative and anti-inflammatory effects[27–29], and could therefore potentially mitigate negative symptoms of schizophrenia and BD. Opposite to the results for anorexia, the MR analysis showed that both pyridoxal and ferulic acid 4-sulfate have protective effects on both BD and schizophrenia, which implies that a dietary supplement of ferulic acid and vitamin $B_6$ could be beneficial for supplementary treatment of the disorders.

cAMP is an adenosine triphosphate (ATP) derivative which serves primarily as a second messenger and has a central role in cellular functions, metabolism, and neurotransmission[30]. cAMP signalling abnormalities have strong associations with schizophrenia[31–33] as well as BD[34–36]. Past studies have shown cAMP differences in blood[37,38] and urine[39,40] of schizophrenia patients, however these studies were conducted nearly half a century ago with different diagnostic criteria, now-outdated measurement methods, inconclusive results, and lower sample sizes. The MR analysis results show that upregulated cAMP concentrations in urine are a risk factor for both schizophrenia and BD, which warrants a reopening of the discussion around cAMP as a potential biomarker of schizophrenia and BD.

### References

1. Wolfsegger, T., Böck, K., Schimetta, W., von Oertzen, T. J. & Assar, H. N,N-Dimethylglycine in patients with progressive multiple sclerosis: result of a pilot double-blind, placebo, controlled randomized clinical trial. *Neurol Res Pract* **3**, 29 (2021).

2. Kern, J. K. *et al.* Effectiveness of N,N-dimethylglycine in autism and pervasive developmental disorder. *J Child Neurol* **16**, 169–173 (2001).
3. Parra, M., Stahl, S. & Hellmann, H. Vitamin B6 and Its Role in Cell Metabolism and Physiology. *Cells* **7**, 84 (2018).
4. Sure, B. A Detailed Study of the Role of Vitamin B in Anorexia in the Albino Rat. *J Nutr* **1**, 49–56 (1928).
5. Rock, C. L. & Vasantharajan, S. Vitamin status of eating disorder patients: Relationship to clinical indices and effect of treatment. *International Journal of Eating Disorders* **18**, 257–262 (1995).
6. Halter, B., Ildari, N., Cline, M. A. & Gilbert, E. R. Ferulic acid, a phytochemical with transient anorexigenic effects in birds. *Comp Biochem Physiol A Mol Integr Physiol* **259**, 111015 (2021).
7. de Melo, T. S. *et al.* Ferulic acid lowers body weight and visceral fat accumulation via modulation of enzymatic, hormonal and inflammatory changes in a mouse model of high-fat diet-induced obesity. *Brazilian Journal of Medical and Biological Research* **50**, (2017).
8. Ree, R., Varland, S. & Arnesen, T. Spotlight on protein N-terminal acetylation. *Exp Mol Med* **50**, 1–13 (2018).
9. Huang, N. *et al.* A pilot case-control study on the association between N-acetyl derivatives in serum and first-episode schizophrenia. *Psychiatry Res* **272**, 36–41 (2019).
10. Molina, V. *et al.* Dorsolateral prefrontal N-acetyl-aspartate concentration in male patients with chronic schizophrenia and with chronic bipolar disorder. *European Psychiatry* **22**, 505–512 (2007).
11. Yoshimi, N. *et al.* Blood metabolomics analysis identifies abnormalities in the citric acid cycle, urea cycle, and amino acid metabolism in bipolar disorder. *BBA Clin* **5**, 151 (2016).
12. Saiz, P. A. *et al.* N-acetyltransferase-2 polymorphisms and schizophrenia. *European Psychiatry* **21**, 333–337 (2006).
13. Luan, Z., Lu, T., Yue, W., Copray, S. & Zhang, D. Association between NAT2 polymorphisms and the risk of schizophrenia in a Northern Chinese Han population. *Psychiatr Genet* **27**, 71–75 (2017).
14. Saleem, S., Shaukat, F., Gul, A., Arooj, M. & Malik, A. Potential role of amino acids in pathogenesis of schizophrenia. *Int J Health Sci (Qassim)* **11**, 63 (2017).
15. Erhardt, S., Schwieler, L., Imbeault, S. & Engberg, G. The kynurenine pathway in schizophrenia and bipolar disorder. *Neuropharmacology* **112**, 297–306 (2017).
16. Abdullina, A. A. *et al.* The neuropeptide cycloprolylglycine produces antidepressant-like effect and enhances BDNF gene expression in the mice cortex. *Journal of Psychopharmacology* **36**, 214–222 (2022).
17. Chiappelli, J. *et al.* Tryptophan Metabolism and White Matter Integrity in Schizophrenia. *Neuropsychopharmacology* **41**, 2587 (2016).
18. Bartoli, F. *et al.* The kynurenine pathway in bipolar disorder: a meta-analysis on the peripheral blood levels of tryptophan and related metabolites. *Mol Psychiatry* **26**, 3419–3429 (2020).

19. He, Y. *et al.* Schizophrenia shows a unique metabolomics signature in plasma. *Transl Psychiatry* **2**, e149–e149 (2012).
20. Mew, N. A. *et al.* Urea Cycle Disorders Overview. *GeneReviews®* (2017).
21. Viegas, C. M. *et al.* Experimental evidence that ornithine and homocitrulline disrupt energy metabolism in brain of young rats. *Brain Res* **1291**, 102–112 (2009).
22. Moutsatsou, P. *et al.* Peripheral blood lymphocytes from patients with bipolar disorder demonstrate apoptosis and differential regulation of advanced glycation end products and s100b. *Clin Chem Lab Med* **52**, 999–1007 (2014).
23. D'cunha, N. M. *et al.* The Effects of Dietary Advanced Glycation End-Products on Neurocognitive and Mental Disorders. *Nutrients* **14**, (2022).
24. Firth, J. *et al.* The effects of vitamin and mineral supplementation on symptoms of schizophrenia: a systematic review and meta-analysis. *Psychol Med* **47**, 1515–1527 (2017).
25. Badrfam, R. *et al.* The efficacy of vitamin B6 as an adjunctive therapy to lithium in improving the symptoms of acute mania in patients with bipolar disorder, type 1; a double-blind, randomized, placebo-controlled, clinical trial. *Brain Behav* **11**, e2394 (2021).
26. Mitchell, E. S., Conus, N. & Kaput, J. B vitamin polymorphisms and behavior: Evidence of associations with neurodevelopment, depression, schizophrenia, bipolar disorder and cognitive decline. *Neurosci Biobehav Rev* **47**, 307–320 (2014).
27. Sasaki, K., Iwata, N., Ferdousi, F. & Isoda, H. Antidepressant-Like Effect of Ferulic Acid via Promotion of Energy Metabolism Activity. *Mol Nutr Food Res* **63**, (2019).
28. Zeni, A. L. B., Camargo, A. & Dalmagro, A. P. Ferulic acid reverses depression-like behavior and oxidative stress induced by chronic corticosterone treatment in mice. *Steroids* **125**, 131–136 (2017).
29. Zeni, A. L. B., Zomkowski, A. D. E., Maraschin, M., Rodrigues, A. L. S. & Tasca, C. I. Ferulic acid exerts antidepressant-like effect in the tail suspension test in mice: Evidence for the involvement of the serotonergic system. *Eur J Pharmacol* **679**, 68–74 (2012).
30. Yamamizu, K. & Yamashita, J. K. Roles of Cyclic Adenosine Monophosphate Signaling in Endothelial Cell Differentiation and Arterial-Venous Specification During Vascular Development. *Circulation Journal* **75**, 253–260 (2011).
31. Funk, A. J., McCullumsmith, R. E., Haroutunian, V. & Meador-Woodruff, J. H. Abnormal Activity of the MAPK- and cAMP-Associated Signaling Pathways in Frontal Cortical Areas in Postmortem Brain in Schizophrenia. *Neuropsychopharmacology* **37**, 896 (2012).
32. Tardito, D. *et al.* Abnormal Levels of cAMP-dependent Protein Kinase Regulatory Subunits in Platelets from Schizophrenic Patients. *Neuropsychopharmacology* **23**, 216–219 (2000).

33. Nishino, N. *et al.* Increase in [3H]cAMP binding sites and decrease in Giα and Goα immunoreactivities in left temporal cortices from patients with schizophrenia. *Brain Res* **615**, 41–49 (1993).
34. Dwivedi, Y. & Pandey, G. N. Adenylyl cyclase-cyclicAMP signaling in mood disorders: Role of the crucial phosphorylating enzyme protein kinase A. *Neuropsychiatr Dis Treat* **4**, 161 (2008).
35. Ren, X. *et al.* Alteration of cyclic-AMP response element binding protein in the postmortem brain of subjects with bipolar disorder and schizophrenia. *J Affect Disord* **152–154**, 326–333 (2014).
36. Perez, J. *et al.* Abnormalities of cyclic adenosine monophosphate signaling in platelets from untreated patients with bipolar disorder. *Arch Gen Psychiatry* **56**, 248–253 (1999).
37. Garver, D. L., Johnson, C. & Kanter, D. R. Schizophrenia and reduced cyclic AMP production: Evidence for the role of receptor-linked events. *Life Sci* **31**, 1987–1992 (1982).
38. Kafka, M., van Kammen, D. P. & Bunney, W. E. Reduced cyclic AMP production in the blood platelets from schizophrenic patients. *https://doi.org/10.1176/ajp.136.5.685* **136**, 685–687 (2006).
39. Brown, B. L., Salway, J. G., Albano, J. D., Hullin, R. P. & Ekins, R. P. Urinary Excretion of Cyclic Amp and Manic-Depressive Psychosis. *The British Journal of Psychiatry* **120**, 405–408 (1972).
40. Paul, M. I., Cramer, H. & Goodwin, F. K. Urinary Cyclic AMP Excretion in Depression and Mania: Effects of Levodopa and Lithium Carbonate. *Arch Gen Psychiatry* **24**, 327–333 (1971).
